# Urban Exposures, Frailty, and Mental Illness in World Trade Center Health Program Responders

**DOI:** 10.64898/2026.01.09.26343773

**Authors:** Helena Krasnov, William Hung, Pablo Knobel, Itai Kloog, Fred Ko, Hannah Thompson, Elena Colicino, Susan L. Teitelbaum, Allan. C Just, Maayan Yitshak Sade

**Author notes:** Corresponding author: Helena Krasnov, Department of Environmental Medicine Icahn School of Medicine at Mount Sinai New York, NY.

## Abstract

General responders of the World Trade Center (WTC) Health Program were uniquely exposed to chemical toxicants and extreme psychological stress during the 2001 terrorist attack, and now experience unusually prevalent health outcomes for a mid-aged population, including frailty, Post Traumatic Stress Disorder (PTSD), and depression. We studied whether these conditions are linked to later-life urban environmental exposures among 18,734 responders, evaluating associations with annual fine particulate matter (PM_2.5_), temperature, and Green View Index. A decile incremental increase in the overall exposure-mixture was associated with increased frailty (0.56%; 95% CI 0.14%;0.98%) and PTSD scores (0.31%; 95% CI 0.07%;0.55%). Effects were more pronounced among responders with higher WTC exposures, suggesting increased vulnerability. Here, we show that urban environmental exposures compound the chronic health burden in WTC responders, with effects possibly amplified by WTC-exposure history. This suggests that a history of trauma and toxicant exposure can create lasting biological vulnerability to subsequent environmental stressors.

## INTRODUCTION

General responders who took part in the rescue, recovery, and cleanup efforts after the terrorist attack on the Twin Towers on September 11, 2001, suffer severe health consequences to this day^1^. These health conditions are monitored as part of the World Trade Center Health Program (WTCHP) ^1^. WTCHP responders were uniquely exposed to a combination of chemical toxicants and extreme psychological stress during and after the World Trade Center (WTC) disaster. Moreover, studies show high rates of post-traumatic stress disorder (PTSD) and mental health conditions among this group ^2,3^. Further, despite the relatively young age, frailty is highly prevalent in this cohort ^4^. Frailty and depression are dynamic conditions that are distinct but interrelated in the aging population ^5^. These conditions share similar symptoms and risk factors, including chronic inflammation, oxidative stress and vascular disease ^5^.

Recent studies show that, beyond historic WTC-related effects, environmental exposures later in life can also shape the risk of various chronic conditions ^6,7^. We focus on three exposures: fine particulate air pollution (PM_2.5_), temperature, and greenness, because they represent three distinct but co-occurring domains of the external environment, each with emerging evidence linking them to the health outcomes of interest in this cohort. In dense urban areas, these exposures are physically linked. The green space acts as a physical shield providing shade ^8^ and “evapotranspiration”, which directly lowers local temperatures. High temperatures act as a catalyst for air pollution. Heat speeds up the chemical reactions that turn tailpipe emissions into ground-level ozone and secondary particulates ^9^. Tall buildings trap heat and pollutants while blocking the cooling effects of small green patches. This creates “micro-environments” where high heat and high pollution coexist ^10^.

Emerging evidence suggests that air pollution exposure ^11,12^, cooler ^13,14^ and hotter temperatures ^15,16^, and greenness ^17^ are each independently associated with frailty and depression. However, evidence on the associations of these environmental risk factors with PTSD are scarce, with existing studies focusing mainly on extreme weather events such as hurricanes and floods ^18–20^. Additionally, studies investigating these exposures simultaneously are limited. Research in military populations ^21^ exposed to acute psychological stress and complex chemical and environmental toxins highlights the importance of investigating environmental determinants of mental health in cohorts with heightened vulnerability and elevated exposure burdens. The WTCHP responder cohort, sharing these defining characteristics, represents a critical population in which to examine these associations.

As this cohort ages and frailty and mental illness become more prevalent, understanding how these exposures jointly shape long-term health risk is increasingly important. Each exposure also offers a unique measurement contribution. In this study, rather than relying solely on satellite-based Normalized Difference Vegetation Index (NDVI), we use Green View Index (GVI), which captures additional dimensions of greenness, such as green volume. This makes GVI a more relevant measure of human-experienced greenness, particularly in dense urban settings ^22,23^. In this work, we examine whether urban environmental exposures in New York City (NYC) during ∼25 years of follow-up (i.e., PM_2.5_, temperature, and greenness exposure) increase the risks of frailty, PTSD scores, and depression scores among WTC general respondents living in NYC during the years 2003-2023. We show that higher PM_2.5_ and temperature exposures are associated with increased risks of frailty, depression, and PTSD, while residential greenness is associated with a significant decrease in these risks. Investigating the overall exposure-mixture effect, we found increases in frailty and PTSD scores associated with incremental increases in the three exposures, simultaneously highlighting the need to consider the complex interplay between environmental factors when investigating their impacts on health. Additionally, we investigate WTC exposure intensity as an effect modifier, with results suggesting that associations may be stronger among responders with greater exposure burden. These findings have significant implications for aging populations, particularly cohorts with heightened environmental vulnerability such as WTCHP responders.

## RESULTS

We included 81,295 frailty index (FI) scores, 115,153 PTSD scores, and 109,220 depression scores of 18,734 responders. About 73% contributed at least four visits (**Figure S1A**), and most had their first visits earlier years (**Figure S1B**). The average ages on the first and last visits were approximately 47 and 58 years. Most responders were White and 81.88% were males (**Table 1**). The average count of deficits FI per individual was 5.80. Specific deficit frequencies are presented in **Table S1**. Approximately 39% met the criterion for probable PTSD and 35% met the criterion for probable depression. The summary statistics for the exposures are presented in **Table 2**. The correlation between the exposures was low, with the highest correlation between PM_2.5_ and GVI (r = 0.04).

**Table 1.**
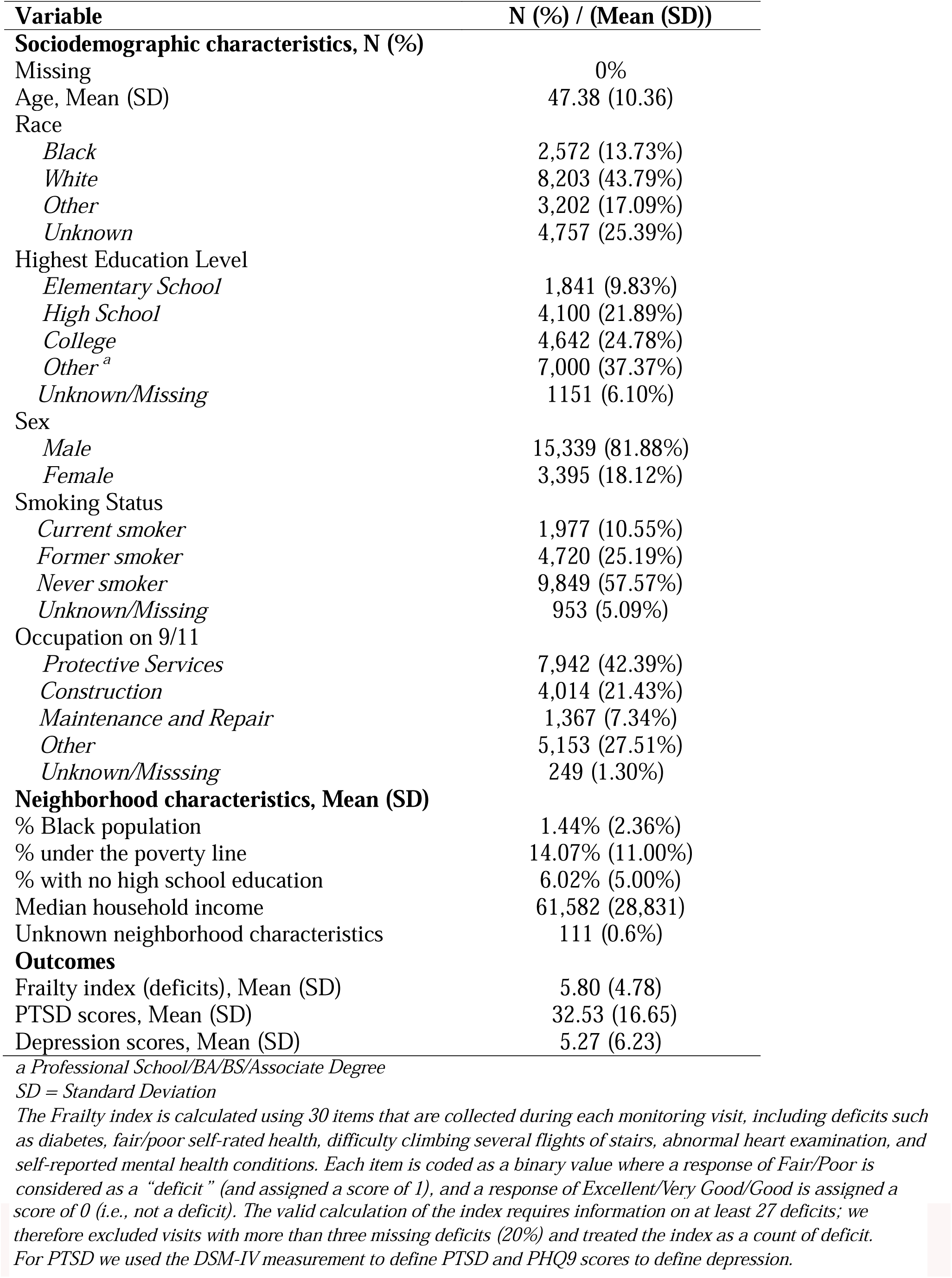
Population characteristics at cohort entry (N = 18,734), 2003-2023.

**Table 2.** Summary statistics of annual exposures and modifiers, 2003-2023.

| Variable | Mean (SD)/ N (%) | IQR |
| --- | --- | --- |
| **Exposures, Mean (SD)** |  |  |
| PM<sub>2.5</sub> (µg m<sup>-3</sup>) | 8.19 (1.74) | 2.48 |
| Mean temperature (° C) | 13.14 (2.12) | 1.32 |
| Green View Index, 300m buffer * | 14.90 (5.30) | 6.67 |
| **Modifiers, N (%)** |  |  |
| WTC exposure index ** |  |  |
| Low/Intermediate | 14,046 (75.00%) |  |
| High/Very High | 3,655 (19.50%) |  |
| Missing | 1,033 (5.50%) |  |
| Witnessed horror |  |  |
| No | 10,537 (56.25%) |  |
| Yes | 6,278 (33.51%) |  |
| Missing | 1,919 (10.20%) |  |
| Arrival date |  |  |
| Arrived on 9/11-9/14 | 13,346 (71.20%) |  |
| Arrived after 9/14 | 5,334 (28.50%) |  |
| Missing | 54 (0.30%) |  |

\* The Green View Index (GVI) measures the percentage of visible tree canopy from panoramic street view images
\*\* The WTC exposure index is a derived exposure variable that incorporates data on the overall time working at the WTC site, exposure to the dust cloud, and direct work on the debris pile. The “low”/“intermediate” category included responders who worked less than 90 days, were not exposed to the dust, and did not work directly on the pile. The “high”/“very high” category included responders who were directly exposed to the cloud or worked for more than 90 days.

First, we used multivariable regressions to assess the associations with the outcome for interquartile range (IQR) incremental increases in the exposures (95% Confidence Interval [CI]). Higher PM_2.5_ exposure was associated increases in FI (2.27% [0.64%, 3.93%]); PTSD (2.12% [1.33%, 2.92%]) and depression (3.92% [1.95%, 5.92%]) scores. Higher mean temperature was also associated with increases in FI (0.83% [0.47%, 1.19%]); PTSD (0.64% [0.46%, 0.82%]), and depression (0.80% [0.36%, 1.25%]) scores. Finally, higher GVI exposure was associated with decreases in FI (-1.43% [-2.19%,-0.67%]), PTSD (-1.85% [-2.24%,-1.46%]), and depression (-3.83% [-4.77%,-2.87%]) scores (**Table 3**).

**Table 3.** The association between environmental exposures and Frailty, PTSD, and depression scores.

| Variable | % Change (95% CI) |  |  |
| --- | --- | --- | --- |
|  | FI | PTSD | Depression |
| **Multivariate model** |  |  |  |
| PM<sub>2.5</sub> (µg m<sup>-3</sup>) | 2.27% (0.64; 3.93) | 2.12% (1.33; 2.92) | 3.92% (1.95; 5.92) |
| Mean temperature (° C) | 0.83% (0.47; 1.19) | 0.64% (0.46; 0.82) | 0.80% (0.36; 1.25) |
| Green View Index, 300m buffer | -1.43% (-2.19; -0.67) | -1.85% (-2.24; -1.46) | -3.83% (-4.77; -2.87) |
| **Exposure-mixture model** |  |  |  |
| Exposure mixture | 0.56% (0.14; 0.98) | 0.31% (0.07; 0.55) | 0.32% (-0.25; 0.90) |

FI = Frailty Index; First, we used multivariate mixed-effect Poisson models to assess the associations of the three exposures with repeated measures of each outcome. Then, we used quantile-based g-Computation models (Qgcomp) to estimate the overall exposure-mixture associations of PM<sub>2.5</sub>, temperature, and Green View Index with each outcome. Models are adjusted for year of testing, self-reported sex, age, race, education, smoking status, occupation on 9/11 and neighborhood sociodemographic variables.

When evaluating the overall exposure-mixture effect, we found a decile incremental increase in the exposure mixture to be associated with a 0.56% (0.14%, 0.98%) increase in FI and 0.31% increase in PTSD score (0.07%, 0.55%). These associations were driven primarily by mean temperature, which contributed 98% of the overall effect for FI and 73% for PTSD scores. The GVI component in both mixtures was negatively associated with the outcome, but the positive effects were stronger, making the overall exposure-mixture effects positive (**Figure 1**). We did not find an association between the exposure mixture and depression test scores (0.32% [-0.25%; 0.90%]). As shown in Table S2 PM_2.5_ and temperature were associated with higher odds for abnormal PTSD and depression scores, while GVI was associated with lower odds of abnormal scores. The overall exposure mixture were 1.01 (1.00; 1.03) for PTSD and 1.01 (0.99; 1.02) for depression (**Table S2**).

**Figure 1.**
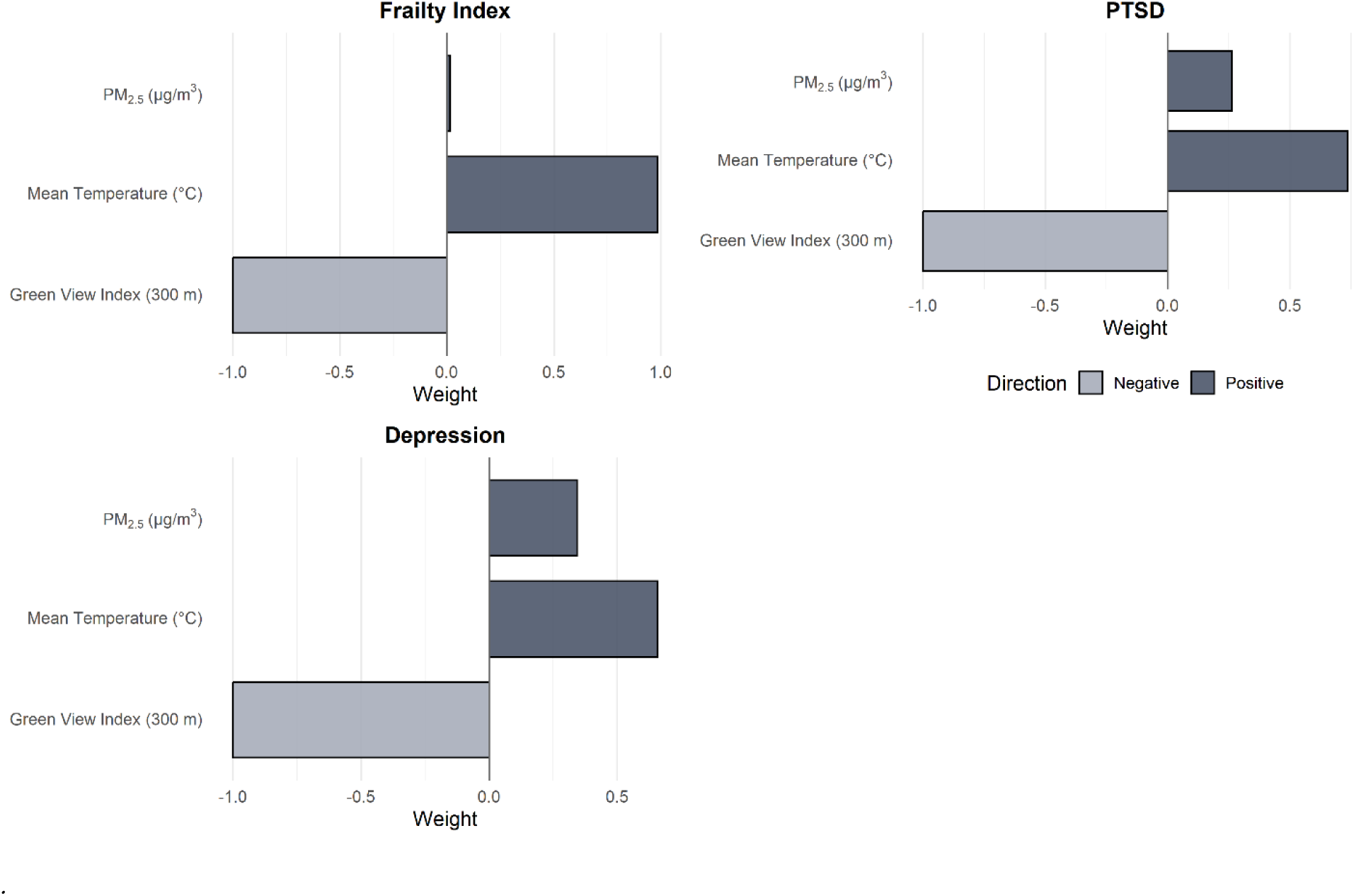
The weighted contribution of the components of the exposure-mixture to the associations with frailty, PTSD, and depression scores.

In a secondary analysis examining interactions between exposure-mixture effects and WTC exposures, associations were consistently larger among responders with higher WTC-exposure index rankings, early site arrival (within the first three days), and those who witnessed horror. However, only the association with depression was statistically significantly modified by witnessing horror (interaction p value 0.04) (**Table 4**).

**Table 4.**
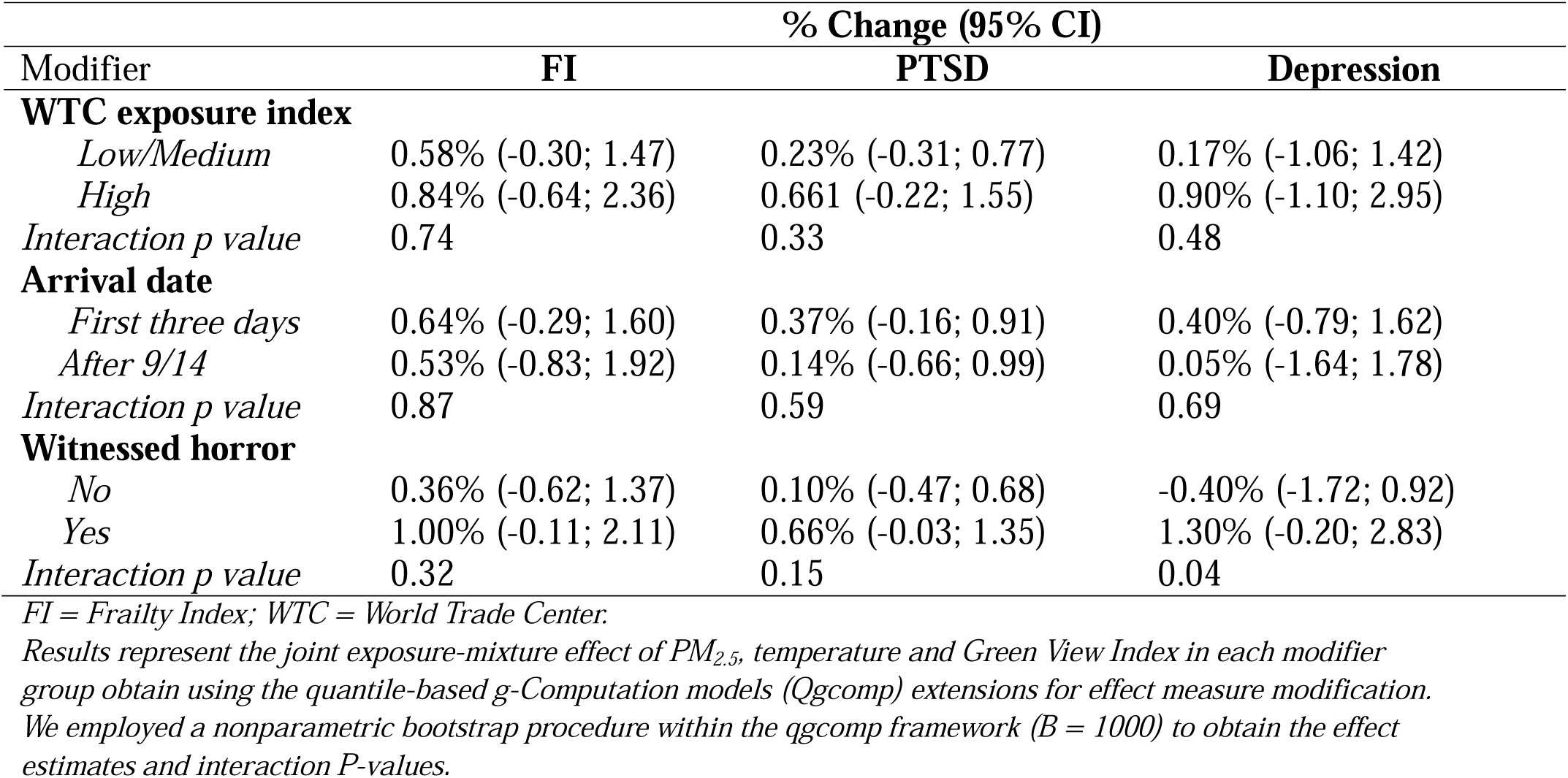
Effect modification of the associations between the exposure-mixture Frailty, PTSD, and depression scores by WTC exposures.

We conducted several sensitivity analyses to evaluate the sensitivity of our results to study assumptions. First, when using 500m buffer to evaluate GVI exposure we find very similar results, although the 300m buffer used in the main analyses shows stronger associations with the outcomes (**Table S3**). Second, we repeated the models assessing the associations with FI while incorporating inverse probability weights to account for the probability of inclusion in the study.

The associations were similar and the overall inference did not change (**Table S4**). Thirdly, we repeated the analysis among a sub sample restricted to observation years 2012 and after.

Although less precise due to smaller sample size, the results were similar and the overall inference did not change (**Table S5**). Finally, we repeated the analysis using summer and winter exposures and found that the overall inference did not change (**Table S6**).

## DISCUSSION

Our results are consistent across diverse model specifications linking environmental exposures to frailty, depression, and PTSD risk among general responders enrolled in the WTCHP. Higher PM_2.5_ and temperature exposures were associated with increased risks of frailty, depression, and PTSD, while residential greenness was associated with a significant decrease in these risks.

When examining the overall exposure-mixture effect, simultaneous increases across all three exposures were associated with higher FI and PTSD scores, with temperature as the dominant driver; suggesting that the harmful effects of temperature and PM_2.5_ outweigh the protective benefit of greenness. While associations with continuous PTSD and depression symptom scores were modest, higher PM_2.5_ and temperature and lower greenness exposures were significantly associated with a higher risk of meeting clinical thresholds for these conditions. Finally, although not statistically significant, the higher effect estimates may suggest that responders with more intense WTC-related exposures may be more vulnerable to the effects of later-life environmental exposure mixtures on aging and mental health.

Evidence of single-exposure associations from environmental, aging, and mental health studies is accumulating ^24^. In accordance with our findings, a 2019 meta-analysis concluded that a10 μg/m^3^ increase in long-term exposure to PM_2.5_ is associated with 10% increased risk of depression ^25^. Studies of PM_2.5_ exposure also found increased odds of frailty associated with higher exposures ^26–28^. These associations may be related to oxidative stress and inflammatory responses ^29^ or disrupted dopamine signaling ^30^. Increased temperature was also associated with elevated risks of frailty and mental illness in our study. These associations were observed in other studies^15,16^, and may be attributed to the higher thermal sensitivity ^31^ and to heat-induced alterations in autonomic function, which may, in turn, trigger an inflammatory reaction ^32^.

Finally, studies link higher greenness exposure with lower levels of depression, stress ^33,34^, and frailty ^35^, potentially attributed to better opportunities for physical activity and social gatherings35-37.

A unique finding of our study is the link between PM_2.5_ exposure and higher PTSD scores. Studies evaluating the association between PM_2.5_ and PTSD are very limited, and most have reported no association ^38,39^. We hypothesize that moderate air pollution exposure does not directly cause PTSD but instead exacerbates pre-existing mental health conditions such as depression. Studies examining the associations with temperature are also scarce. A few studies found an increase in daily emergency psychiatric visits associated with exposure to hotter temperatures ^40,41^. Excessive heat may affect the course of mental illness through altered thermoregulation and impaired behavioral adaptations, thereby disrupting sleep, recreation, self-care, and social interaction routines, which can lead to increased fatigue and irritability ^41^.

Another important finding is the evidence of the overall exposure-mixture associations. Considering the inherent co-exposure among these variables, we treated the three exposures as a mixture to explicitly model their combined association with frailty and mental health risks. We found that higher PM_2.5_ and temperature exposures were associated with elevated frailty and PTSD risks, while accounting for the mitigating of greenness. Temperature emerged as the primary driver of exposure-mixture associations, likely reflecting heat island effects common in dense urban areas like NYC. Unlike localized stressors such as noise and air pollution, the Urban heat island effect imposes a long and persistent physiological strain that is harder for vulnerable populations to mitigate ^42^. Our sensitivity analysis using summer and winter temperature exposure provides further support that hotter temperatures are associated with larger risks of the outcomes. To our knowledge, there are no other studies examining the associations between this urban exposure mixture and frailty or PTSD. Other studies, however, investigated other combinations of exposure mixtures and related mental health outcomes ^43–45^, and linked environmental exposure-mixtures to depressive symptoms ^43^, general anxiety disorder ^44^, and frailty ^45^.

An emerging finding of our study is that associations between the exposure mixture and mental health outcomes were amplified among responders with more intense WTC-related exposures. This was most pronounced for depression, where significantly larger mixture associations were observed among those who witnessed horror during the 2001 attack. Although not reaching statistical significance, similar patterns of amplified associations across depression, frailty, and PTSD were observed among those assigned to higher WTC exposure index categories and those who arrived at the site within the first three days. These results extend the well-documented link between trauma and mental illness ^46^ reinforcing previously reported connections between WTC exposure severity, PTSD, and depression ^47^. The biological mechanisms contributing to these health effects are not fully understood, but some studies report on damaged neural mechanisms in the WTC cohort due to the WTC exposures ^48^. These results support a’two-hit’ model of health impairment: initial WTC exposures may prime the body for increased sensitivity to subsequent environmental hazards. Specifically, we observed that higher WTC exposure intensity exacerbated the impact of air pollution, similar to previously reported interactions affecting metabolic health markers within this population ^7^. This hypothesis is supported by evidence from a separate study showing that individuals experiencing a higher frequency of extreme environmental disasters (e.g., annual wildfire counts) were more susceptible to the health effects of all-source air pollution ^49^. Given the lack of statistical significance in most our interaction models, future studies are required to further explore this possible relationship.

Our study has several limitations. Firstly, the GVI exposure was measured cross-sectionally. While this limits our ability to assess time-varying effects, green infrastructure changes slowly over time and therefore can be treated as time-stable. Second, although we adjusted for a robust set of individual and neighborhood-level sociodemographic covariates, residual confounding from unmeasured behavioral or environmental factors remains possible. For example, since complete longitudinal occupational exposure data is not available, we cannot account for these potential confounders in our analysis. However, to minimize confounding bias, we account for occupation prior to 9/11, which is likely to represent occupational exposure in later life.

Additionally, the use of detailed health information and neighborhood data reduces the likelihood that major confounders were omitted and helps strengthen the internal validity of our findings.

Thirdly, while our population is restricted to NYC, it is a unique urban setting with distinctive spatial and social structures that offer valuable insight into how diverse urban nature configurations relate to aging and mental health outcomes. Finally, a consideration of this study is the use of an open cohort design. Firstly, individuals who join the cohort at later years may differ systematically from those who joined in earlier years, potentially introducing selection bias. However, our sensitivity analysis restricting the sample to cohort years following 2012 shows similar results and the overall inference did not change Second, varying follow-up durations across participants can complicate the accurate characterization of long-term or cumulative environmental exposures. To address this issue, we assign time-varying exposure and confounders. Finally, cohort attrition may bias effect estimates in open cohorts. However, since attrition is unlikely to be related to the ambient exposures assessed in our study it unlikely to bias our results.

In conclusion, higher PM_2.5_ and temperature exposures and lower greenness were found to be associated with elevated risks of frailty and worsening depression and PTSD test scores. Further, the overall exposure-mixture association with frailty and PTSD was mainly driven by temperature exposure. While associations with continuous PTSD and depression symptom scores were modest, higher PM_2.5_ and temperature and lower greenness exposures were significantly associated with a higher risk of meeting clinical thresholds for these conditions. Finally, responders who experienced more intense WTC-related exposures might be more vulnerable to later-in-life environmental exposure-mixture effects on mental health and aging outcomes. This study highlights the need to consider the complex interplay between environmental factors when investigating their impacts on health. These findings have significant implications for aging populations, particularly cohorts with heightened environmental vulnerability such as WTCHP responders.

## METHODS

### Study population

We included general responders from the WTCHP cohort, an open cohort that continuously enrolls responders. Eligible individuals include responders who took part in rescue, recovery, and clean-up tasks following the 9/11/2001 attack on the WTC. Complete details regarding study eligibility criteria are described elsewhere and include occupational roles and the total duration of service on-site ^50^. Our initial analytical sample included 46,535 responders who were enrolled between 2001 and 2021. To align with the availability of exposure data, we included only study visits between the years 2003 and 2023. General responders enrolled in the WTCHP are offered annual clinical visits, termed monitoring visits ^50^, during which a healthcare professional performs physical and mental examinations along with a survey of demographic and health-related questions. Data collection occurred during attended visits; at each encounter, respondents were screened for various conditions, laboratory tests were performed, and overall health was evaluated, resulting in the creation of longitudinal cohort with observations for each visit.

Sociodemographic information was collected at the first visit, and address histories were updated at each monitoring visit starting in 2012.

We restricted the population to responders living in NYC (N=18,734) to capture associations in a dense metropolitan environment using high-resolution measure of street-level greenery. A flowchart diagram with numbers of individuals based on certain criteria is presented in **Figure S2**. The cumulative and absolute frequency of visits as well as the frequencies of study visits, are shown in **Figure S1A** and **B** and the distribution of the responders’ (person years) locations is in **Figure S3**.

### Outcomes

Frailty is commonly estimated using a Frailty Index (FI), a measure of age-associated declines across multiple health domains ^51–54^. We used the FI developed specifically for the WTCHP general responder cohort ^51,55^. The FI was constructed from 30 items collected during monitoring visits. Each item was dichotomized: responses of’Fair’ or’Poor’ were classified as deficits, while responses of’Excellent,’’Very Good,’ or’Good’ were classified as non-deficits. A participant’s frailty score at each visit was calculated as the total number of deficits endorsed (range: 0–30).

The valid calculation of the index requires information on at least 27 deficits; we therefore excluded visits with more than three missing deficits (20%). To allow compatibility with the exposure-mixture approach while accounting for clustering of repeated measures within person, we treated the FI as a count of deficits and incorporated the total number of available deficit assessments in the models ^35^.

We determined the continuous PTSD and depression status based on repeated test scores from validated tools administered during the monitoring visits. For PTSD, we used the PTSD DSM-IV measures (PCL), ranging between 17-85. For depression, we used the Patient Health Questionnaire-9 (PHQ-9), ranging between 0-27. As secondary outcomes, we defined PTSD given PCL scores ≥ 44 ^56^ and depression given a score > 10 ^57^. A histogram illustrating the distribution of the studied outcome is presented in **Figure S4**.

### Exposures

We obtained daily PM_2.5_ exposures from geospatial model. The model combines extreme gradient boosting (XGBoost) with satellite-based data in order to predict daily PM_2.5_ on a 1 × 1-km^2^ resolution covering the contiguous United States ^58^. The Model performance was evaluated against PM_2.5_ measurements from monitoring sites, showing a root mean square error of 3.11 μg m-3, which falls in the range of other predictive models ranging between 2-5 μg m-3 ^59^. We obtained temperature exposures from a similar prediction model using XGBoost and inverse distance weighting. The model was evaluated with station-level cross-validation to predict daily temperatures with a root mean square error of 1.19K ^60^. We aggregated the daily exposures to create annual exposures. If a responder moved mid-year, the average value of both locations was used. We calculated the mean GVI at a 300m buffer for the main analysis, and 500m buffer for the sensitivity analysis, around the residential addresses to fully capture the greenness surrounding homes, as commonly done in other studies ^61^. GVI captures the urban street greenery (i.e., tree visibility from the street) using 2020 street view imagery from the Treepedia dataset. Further details can be found in Li et al., 2015 ^62^. Given the limited temporal variability in residential greenness in NYC, it is treated as a proxy for exposure over the entire study period ^63^. All the exposures were linked to each participant’s residential address at the time of the corresponding visit. Since address histories were updated at each monitoring visit starting 2012, for visits occurring before 2012, the address reported at the 2012 visit was used.

### Covariates and modifiers

Models were adjusted for individual-and census tract-level characteristics. Individual-level covariates included age (time-varying), self-reported sex, race (Black, White, other, or unknown), education (elementary school, high school, college, or other/unknown), smoking status (time-varying; currently smoking or former or non-smoker), and occupation during 9/11 (construction; protective services; maintenance and repair; or other). Census tract-level socioeconomic variables were also updated at each visit to reflect participants’ residential location over time, and included percentage of residents below the poverty line, percentage of Black residents, percentage without a high school diploma, and median household income.

As a possible modifiers, we selected environmental and psychological WTC exposures previously related to chronic health outcomes among the responders ^6,64^. We gathered the following information from the exposure assessment questionnaire administered during the first monitoring visit: WTC exposure index (low-intermediate/high-very high), arrival date (9/11-13 or 9/14 and after) and whether the responder witnessed horror (yes/no). The WTC exposure index is a derived exposure variable that incorporates data on the overall time working at the WTC site, exposure to the dust cloud, and direct work on the debris pile ^47^.

## Statistical analysis

We used multivariable generalized additive mixed-effect models to simultaneously assess the associations between the three exposures (i.e., PM_2.5_, temperature, and GVI) and repeated measures of FI, PTSD, and depression scores. For all three outcomes we used a quasi-Poisson link function to account for the over-dispersion ^65^. For FI, the number of variables available for the frailty calculation was used as an offset. We adjusted the models for the covariates described above and used a penalized spline for the year of testing to account for the time trend.

Furthermore, we included random intercept to account for repeated measures within participants. To reduce the influence of extreme values, the upper and lower 0.5% of observations for PM_2.5_ and temperature were excluded. Results are presented for IQR incremental increases in the exposures.

Then, we used quantile-based g-Computation models (Qgcomp) to evaluate the overall exposure-mixture associations with repeated measures of FI, PTSD and depression scores, as well as the relative contribution of each exposure to the overall associations ^66^. Qgcomp provides a flexible framework for estimating the total exposure effect without assuming directional homogeneity across exposures, as defined by the following formula: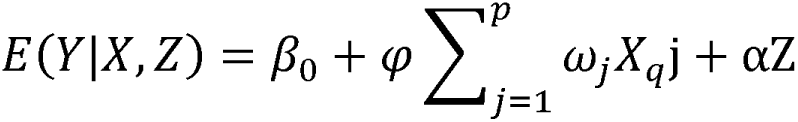 Where: X represents exposure; ψ is the overall mixture effect; w is the weights assigned to each exposure (summing to 1 for positive effects and-1 for negative effects), and Z represents the vector of covariates. We selected deciles based on the lowest Bayesian Information Criterion values after comparing the results of 5 to 10 increments.

Finally, we used the Qgcomp extension for effect measure modification ^67^ to assess differential vulnerability to later-in-life environmental exposures among responders exposed to higher WTC-exposures compared to those with lower exposure levels. We used Poisson regression and employed a nonparametric bootstrap procedure (B = 1000) to obtain robust standard errors, confidence intervals, and interaction P-values.

### Sensitivity analyses

We performed three sensitivity analyses to assess the robustness of our findings. Firstly, since 20% of the visits had fewer than 27 items for the FI calculation, we conducted a sensitivity analysis to account for potential selection bias. We estimated the probability of having at least 27 deficits using a logistic regression model incorporating the following predictors: age, self-reported sex, race, education, smoking status, and occupation during the 9/11, as well as census tract level socioeconomic variables. We repeated the main analyses while incorporating the calculated inverse probability weights (IPW) to assess the robustness of our primary estimates to potential selection bias. Secondly, to ensure our results were not biased due to measurement error, we conducted a sensitivity analysis restricting the data to 2012–2023, during which address histories were collected. Thirdly, to account for temperature variability within a year we included analysis with summer and winter temperature exposures. Due to high correlation (r=0.834) we estimated the associations with each temperature metric separately.

## ETHICAL APPROVAL

This study was approved by the Mount Sinai School of Medicine institutional review board (STUDY-20-02018-MOD004), and consent for the secondary analysis was waived. We restricted the population to responders who consented to data aggregation for research (STUDY-11-02030, World Trade Center Health Program General Responder Data Center).

## DATA AVAILABILITY

The WTC cohort data were obtained from the World Trade Center Health Program (WTCHP) General Responder Cohort (GRC) data center and contain protected health information. Data sharing is restricted under our data use agreement, which prohibits reuse or redisclosure of data obtained under IRB approval. Investigators interested in accessing these data should contact the WTCHP GRC data center directly (https://www.cdc.gov/wtc/researchoutreach.html).

## CODE AVAILABILITY

All code used for data processing and statistical analyses is publicly available on GitHub at [https://github.com/MaayanYSlab/Publications_MaayanYSlab/tree/main/Krasnov2026_Nature% 20Communications].

## Data Availability

The WTC cohort data was obtained from the World Trade Center Health Program (WTCHP) General Responder Cohort (GRC) data center. It includes protected health information with identifiers of individual patients. Per our data use agreement, data supplied for projects with IRB approval shall not be reused or redisclosed. Investigators wishing to request data can contact the WTCHP GRC data center directly.

## AUTHOR CONTRIBUTIONS

Helena Krasnov - Formal analysis, Investigation, Writing - original draft; William Hung - Conceptualization, Methodology, Writing - review & editing; Pablo Knobel - Methodology, Writing - review & editing; Itai Kloog - Software, Validation, Writing - review & editing; Fred Co - Conceptualization, Methodology, Writing - review & editing; Hannah Thompson - Conceptualization, Methodology, Writing - review & editing; Elena Colicino – Statistical analysis and review, Writing - review & editing; Susan L. Teitelbaum - Conceptualization, Methodology, Writing - review & editing; Allan. C, Just - Software, Validation, Writing - review & editing; Maayan Yitshak-Sade - Conceptualization, Methodology, Project administration, Resources, Supervision, Writing - review & editing

## Abbreviations and Acronyms

PM_2.5_: Fine Particulate Matter
WTCHP: World Trade Center Health Program
GRC: General Responders Cohort
WTC: World Trade Center
CVD: Cardiovascular Disease
CI: Confidence Intervals
IQR: Interquartile Range
IPW: Inverse Probability Weights
OR: Odds Ratio
PTSD: Post-Traumatic Stress Disorder
FI: Frailty Index
NYC: New York City
GVI: Green View Index
Qgcomp: Quantile-based g-Computation

## SUPPLEMENTARY INFORMATION

**Figure S1.**
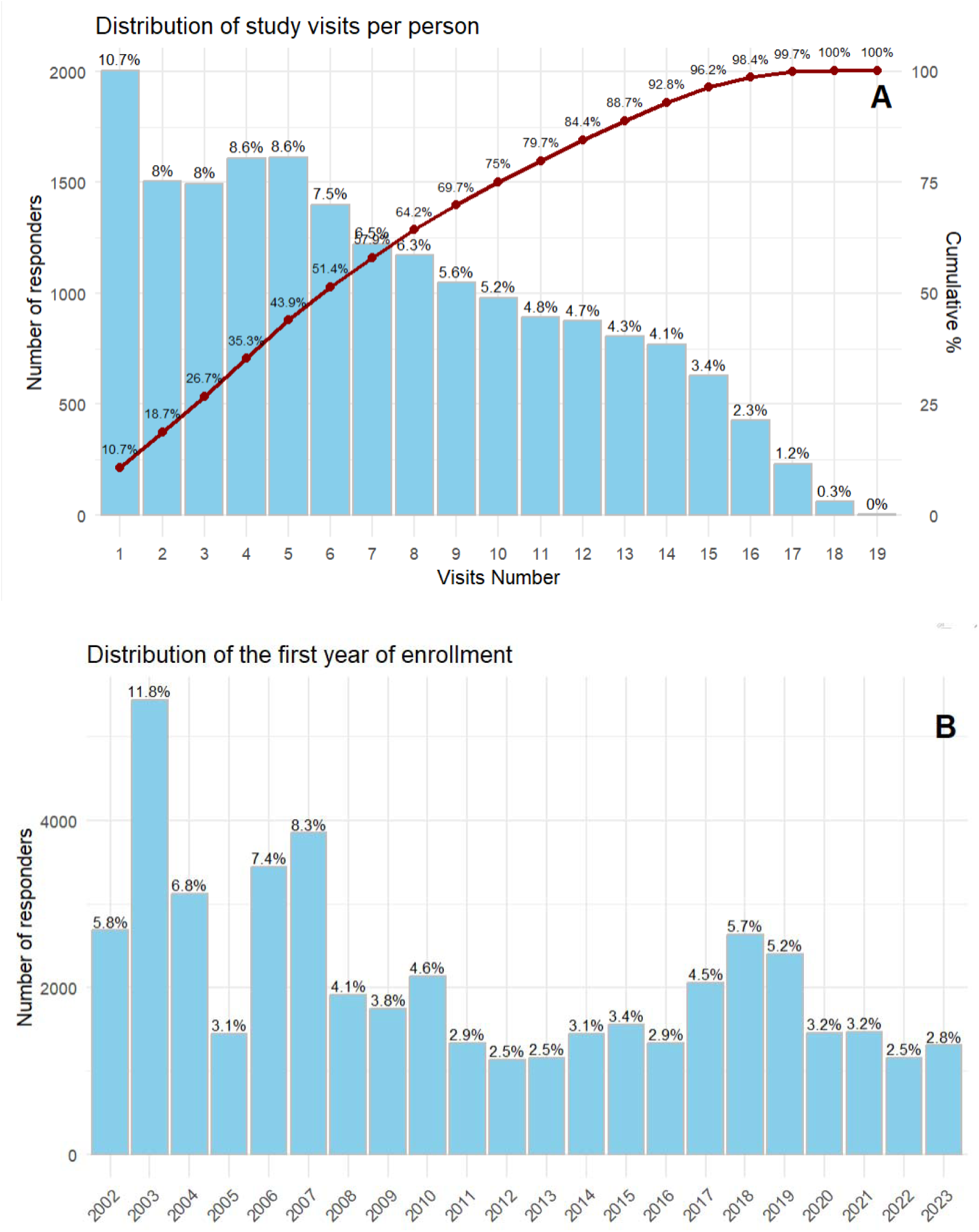
Distribution of the responders monitoring visits

**Figure S2.**
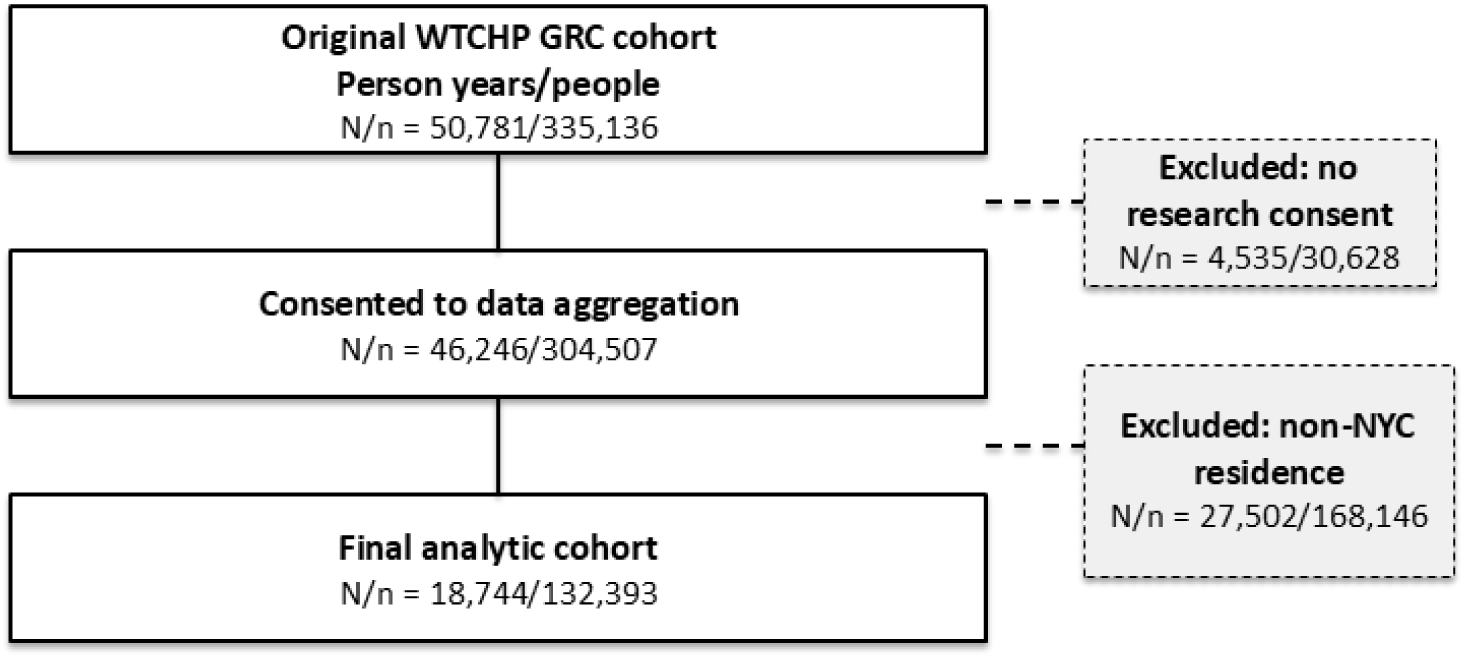
Flowchart of the study population

**Figure S3.**
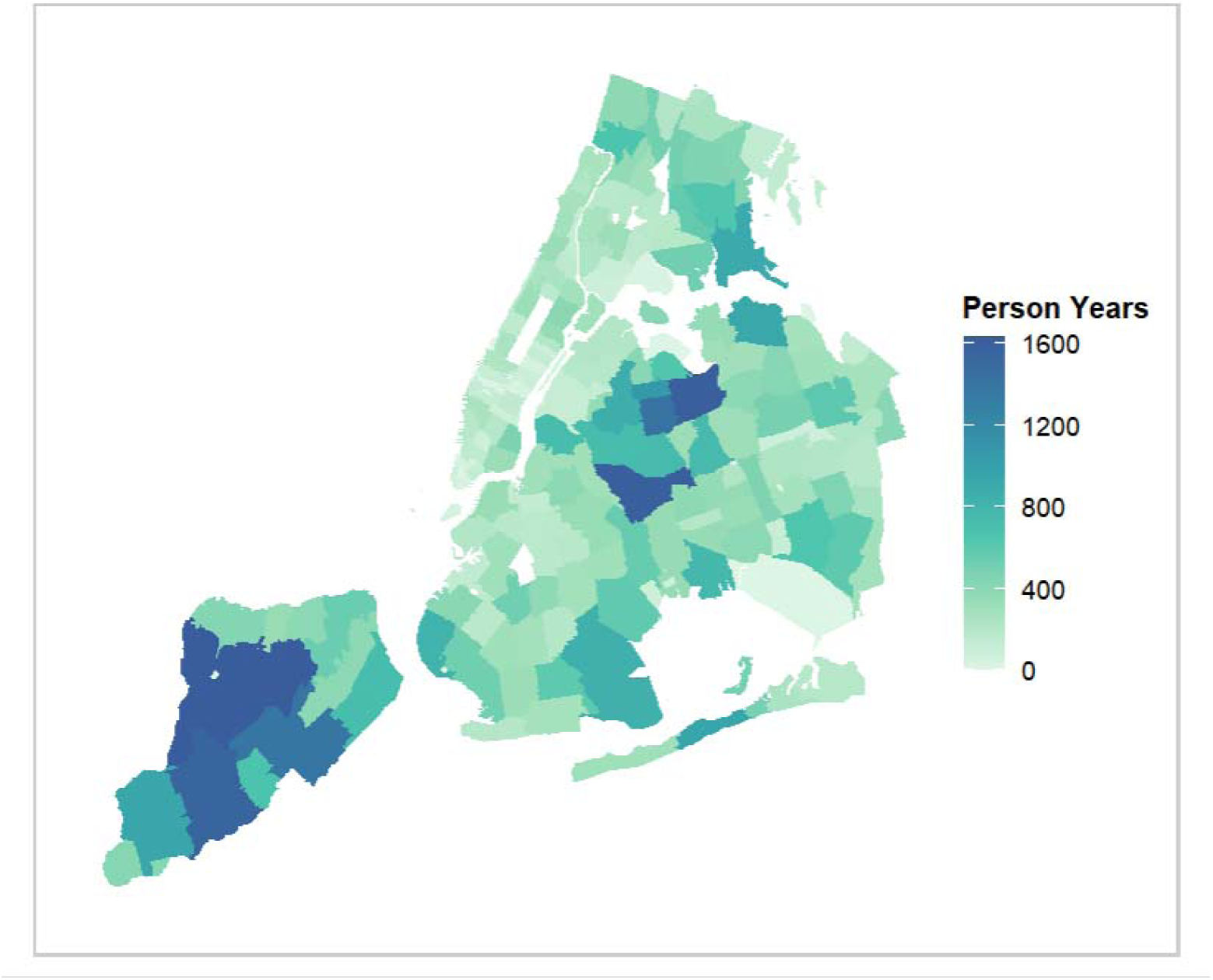
Distribution of the responders’ (person years) locations

**Figure S4.**
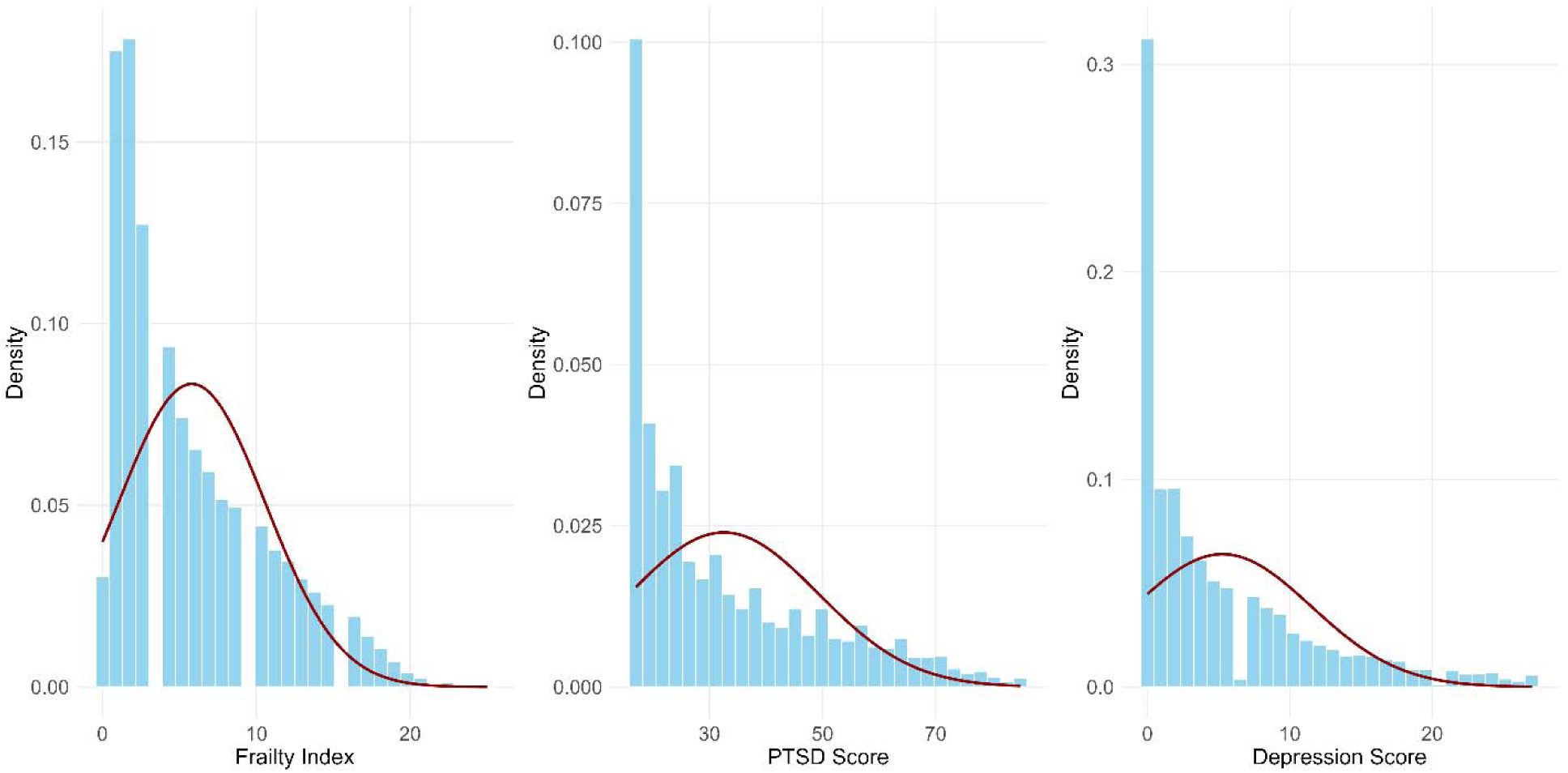
Distributions of the studied outcomes

---

**Table S1.** Number of deficits (person years) between 2003-2023 (N = 134,229)

| Item | Number of<br>deficits<br>(person years) |
| --- | --- |
| Ever had pneumonia | 2 |
| Ever had diabetes | 108 |
| Persistent fatigue | 30,896 |
| Unexplained weight loss | 4,107 |
| Difficulty urinating | 20,159 |
| Inability to taste | 8,227 |
| Difficulty swallowing | 13,547 |
| Change in bowel habits | 15,138 |
| Coughing after you lie down or eat | 16,352 |
| Difficulty hearing | 22,461 |
| Concurrent medications (polypharmacy) | 55,919 |
| Self-rated health | 36,916 |
| Difficulty performing moderate activities (e.g., moving a table, pushing a vacuum cleaner, bowling, or playing golf) | 16,453 |
| Difficulty climbing several flights of stairs | 23,058 |
| In past month, accomplished less than you would like | 44,477 |
| In past month, were limited in the kind of work or other activities you could perform | 44,477 |
| In past month, accomplished less due to emotional problems | 34,087 |
| In past month, did work less carefully than usual due to emotional problems | 25,967 |
| How much does pain interfere with normal work | 23,319 |
| In past month, how often felt calm/peaceful | 14,394 |
| In past month, how often felt lots of energy | 22,410 |
| In past month, how often felt downhearted | 8,620 |
| In the past month, how often have physical health/emotional problems interfered with social activities | 15,635 |
| Frequency of short-term memory problems (e.g., forgetting keys) | 20,877 |
| In past month, frequency of difficulty concentrating | 16,893 |
| Abnormal eye exam | 2,023 |
| Abnormal ear exam | 5,492 |
| Abnormal extremities exam | 6,086 |
| Abnormal heart exam | 3,252 |
| Abnormal physician-assessed general appearance | 85,098 |

*During each monitoring visit, including deficits such as diabetes, fair/poor self-rated health, difficulty climbing several flights of stairs, abnormal heart examination, and self-reported mental health conditions. Each item is coded as a binary value where a response of Fair/Poor is considered as a “deficit” (and assigned a score of 1), and a response of Excellent/Very Good/Good is assigned a score of 0 (i.e., not a deficit)*

**Table S2.** The association between environmental exposures and PTSD, and depression scores: Sensitivity analysis using dichotomous cutoffs.

| Variable | Odds Ratio (95% confidence intervals) |  |
| --- | --- | --- |
|  | PTSD (OR) | Depression (OR) |
| **Multivariate model** |  |  |
| PM<sub>2.5</sub> (µg m<sup>-3</sup>) | 1.12 (1.08; 1.17) | 1.07 (1.02; 1.12) |
| Mean temperature (° C) | 1.02 (1.01; 1.03) | 1.02 (1.01; 1.03) |
| Green View Index, 300m buffer | 0.92 (0.90; 0.94) | 0.93 (0.91; 0.95) |
| **Exposure-mixture model** |  |  |
| Exposure mixture | 1.01 (1.00; 1.03) | 1.01 (0.99; 1.02) |

OR = Odds Ratio

**Table S3.** The association between environmental exposures and Frailty, PTSD, and depression scores using Green View Index with 500m buffer.

| Variable | % Change (95% CI) |  |  |
| --- | --- | --- | --- |
|  | FI | PTSD | Depression |
| **Multivariate model** |  |  |  |
| PM<sub>2.5</sub> (µg m<sup>-3</sup>) | 2.28% (0.68; 3.91) | 2.14% (1.36; 2.92) | 4.19% (2.25; 6.16) |
| Mean temperature (° C) | 0.77% (0.42; 1.12) | 0.60% (0.42; 0.78) | 0.72% (0.29; 1.15) |
| Green View Index, 500m buffer | -1.85% (-2.61; -1.08) | -0.91% (-2.30; -1.15) | -4.40% (-5.36; -3.44) |
| **Exposure-mixture model** |  |  |  |
| Exposure mixture | 0.41% (0.00; 0.82) | 0.22% (-0.00; 0.45) | 0.11% (-0.44; 0.68) |

FI = Frailty Index

**Table S4.**
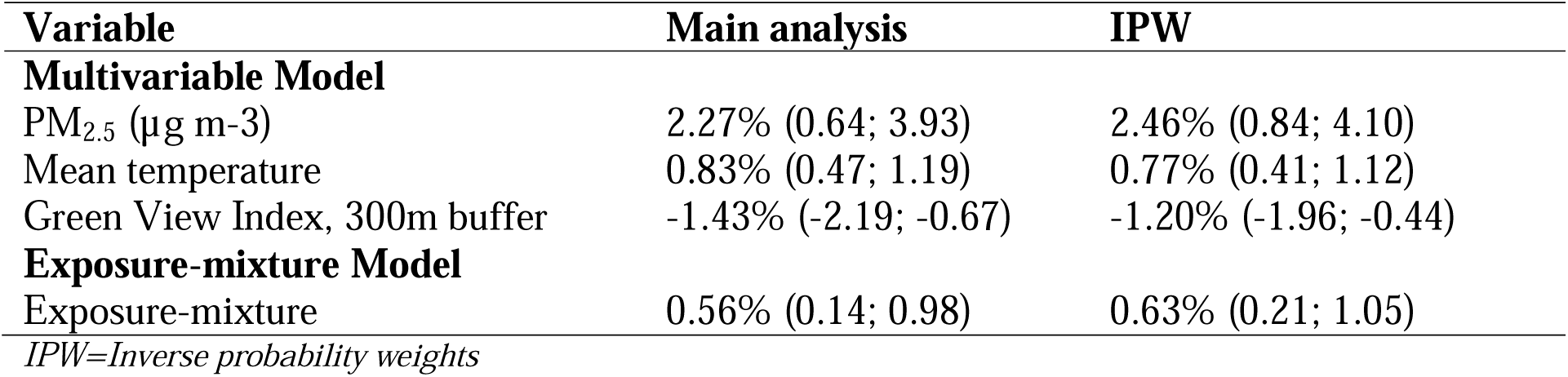
The association between the exposures and Frailty: Sensitivity analysis incorporating the probability of having at least 27 items for the index calculation.

**Table S5.** Sensitivity analysis of the association between environmental exposures and PTSD, restricting the data to 2012–2023.

| Variable | FI | PTSD | Depression |
| --- | --- | --- | --- |
| **Multivariate model** |  |  |  |
| PM<sub>2.5</sub> (µg m<sup>-3</sup>) | 1.87% (-0.18; 3.96) | 1.35% (0.21; 2.50) | 3.11% (0.33; 5.96) |
| Mean temperature (° C) | 0.90% (0.48; 1.31) | 0.70% (0.47; 0.93) | 0.77% (0.21; 1.33) |
| Green View Index, 500m buffer | -1.21% (-2.04; -0.36) | -1.62% (-2.07; -1.16) | -3.29% (-4.39; -2.17) |
| **Exposure-mixture model** |  |  |  |
| Exposure mixture | 0.64% (0.21; 1.07) | 0.28% (0.04; 0.52) | 0.17% (-0.40; 0.75) |
| *FI = Frailty Index* |  |  |  |

**Table S6.** The association between environmental exposures and Frailty, PTSD, and depression scores using different temperature metrices.

| Variable | FI | % Change (95% CI)<br>PTSD | Depression |
| --- | --- | --- | --- |
| **Multivariate model** |  |  |  |
| Summer temperature (° C) | 1.49 % (0.85; 2.14) | 1.29% (0.96; 1.62) | 1.69% (0.90; 2.49) |
| Winter temperature (° C) | 0.66% (0.18; 1.13) | 0.59 (0.34; 0.84) | 0.65% (0.05; 1.24) |

